# Ophthalmic Manifestations of NAA10-Related and NAA15-Related Neurodevelopmental Syndrome: Analysis of Cortical Visual Impairment and Refractive Errors

**DOI:** 10.1101/2024.02.01.24302161

**Authors:** Rahi Patel, Agnes Y. Park, Elaine Marchi, Andrea L. Gropman, Matthew T. Whitehead, Gholson J. Lyon

## Abstract

*NAA10*-related and *NAA15*-related neurodevelopmental syndrome, otherwise known as Ogden Syndrome, is known to present with varying degrees of intellectual disability, hypotonia, congenital cardiac abnormalities, seizures, and delayed speech and motor development. However, the ophthalmic manifestations of *NAA10* and *NAA15* mutations are not yet fully characterized or understood. This study analyzed the prevalence of six ophthalmic conditions (cortical visual impairment, myopia, hyperopia, strabismus, nystagmus, and astigmatism) in 67 patients with pathogenic mutations in the *NAA10* cohort (54 inherited, 10 de novo; 65 missense, 2 frameshift) and 19 patients with pathogenic mutations in the *NAA15* cohort (18 de novo; 8 frameshift, 4 missense, 4 nonsense, and 1 splice site). Patients were interviewed virtually or in-person to collect a comprehensive medical history verified by medical records. These records were then analyzed to calculate the prevalence of these ophthalmic manifestations in each cohort. Analysis revealed a higher prevalence of ophthalmic conditions in our *NAA10* cohort compared to existing literature (myopia 25.4% vs. 4.7%; astigmatism 37.3% vs. 13.2%; strabismus 28.4% vs. 3.8%; CVI 22.4% vs. 8.5%, respectively). No statistically significant differences were identified between the *NAA10* and *NAA15* mutations. Our study includes novel neuroimaging of 13 *NAA10* and 5 *NAA15* probands, which provides no clear correlation between globe size and severity of comorbid ophthalmic disease. Finally, anecdotal evidence was compiled to underscore the importance of early ophthalmologic evaluations and therapeutic interventions.

## Introduction

N-alpha-acetylation is one of the most common eukaryotic post-translational protein modifications. This process is partially facilitated by the NatA complex, which is composed of a N-acetyltransferase 10 (NAA10) catalytic subunit and the NAA15 accessory protein^1–3^. Ogden syndrome (OS), also known as *NAA10*-related and *NAA15*-related syndrome, was first characterized in 2011 as a pathogenic variant in the *NAA10* gene on the X-chromosome^4,5^. Since then, there have been other *NAA10* and *NAA15* variants that have been identified with their phenotypic manifestations, including variable degrees of intellectual disability, hypotonia, congenital cardiac abnormalities, seizures, and delayed speech and motor development^6–16^. A large subset of individuals with *NAA10,* and to a lesser extent *NAA15,* mutations also present with visual abnormalities, including myopia, astigmatism, and cortical visual impairment (CVI)^7^. However, knowledge of the full extent of these ocular manifestations is relatively limited. A deeper understanding of the ophthalmic implications of these mutations is necessary to improve the clinical management of affected patients.

The aims of this study are to describe the ophthalmic manifestations of mutations in *NAA10* in order to compare it to previous findings in the field and to characterize the ophthalmic manifestations of *NAA15* mutations for the first time. We also sought to identify patterns on neuroimaging to correlate globe size to the severity of comorbid ophthalmic pathologies.

## Methods

67 patients (56 females, 11 males) with a pathogenic mutation in *NAA10* and 19 individuals (5 females, 14 males) with a pathogenic mutation in *NAA15* were interviewed via secure video conferencing platforms or in-person at the Jervis Clinic between November 2019 and September 2022. A family of 3 patients with Met147Thr mutations in NAA10 (OS_181, OS_187, OS_188) were not interviewed, but their medical records were provided and sufficiently thorough to include in our analysis. Thorough medical histories were collected, including genetic testing results, information on vision problems, and previous ophthalmic diagnoses. Interviews were 1-2 hours in length and information on all organ systems, as well as social, cognitive, and developmental history, was gathered. Written consent was obtained for the collection of medical records. Google Sheets was used to organize and analyze prevalence data of ophthalmic conditions reported by interviewed participants. The most relevant conditions in patients with *NAA10* and *NAA15* mutations were cortical visual impairment, myopia, hyperopia, astigmatism, strabismus, and nystagmus. Other conditions were reported within the cohort, including anisometropia, eyelid myoclonus, glaucoma, and ptosis. These were not further analyzed due to low prevalence in the cohort and in previous literature.

A PubMed search was conducted to find the prevalence of cortical visual impairment, myopia, hyperopia, strabismus, astigmatism, and nystagmus in previously reported cases of Ogden Syndrome. Initial search criteria included the keywords “Ogden Syndrome” OR “NAA10” AND “ophthalmic OR vision OR visual”; and “Ogden Syndrome” OR “NAA10” AND “cortical visual impairment OR myopia OR hyperopia OR nystagmus OR astigmatism OR strabismus” to gather literature on known reports of visual impairment in patients with NAA10 mutations. Data from the resultant relevant papers (n=106 probands) were organized and compared to data within our cohort. There was no pre-existing literature on the ophthalmic manifestations of *NAA15*, so our findings could only be compared to *NAA10*.

Neuroimaging analysis was also conducted on 18 patients (13 *NAA10* and 5 *NAA15* probands) to assess structural abnormalities of the ocular globes, if present. These findings were analyzed to identify patterns between globe size, globe shape, and ophthalmic diagnoses.

Additionally, to better understand the direct impacts of early ophthalmic intervention, we collected quotes from patient families via email. Some quotes were edited by the authors due to some families not speaking English as their primary language. In these instances, the modified quotes were sent to the families for approval and subsequently approved. Original quotes are included in **Supplementary Table S1.**

## Results

This study includes 67 individuals with *NAA10* variants and 19 individuals with *NAA15* variants. *NAA10* individuals come from 14 countries with a mean age of 10.8 years. *NAA15* individuals come from 4 countries with a mean age of 8.9 years. Within our cohorts, 61 *NAA10* patients (91.0%) and 18 *NAA15* patients (94.7%) were Caucasian. 57 *NAA10* patients (85.1%) and 18 *NAA15* patients (94.7%) were under the age of 18. **Table 1** and **Table 2** outline the demographic information of interviewed patients with *NAA10* and *NAA15* mutations, respectively.

**Table 1.** *NAA10* demographic information of interviewed patients.

| Characteristics | Study Participants (n=67) n (%) |
| --- | --- |
| Sex |  |
| Male | 11 (16.4%) |
| Female | 56 (83.6%) |
| Race |  |
| Caucasian | 61 (91.0%) |
| Asian | 5 (7.5%) |
| African American | 1 (1.5%) |
| Ethnicity |  |
| Hispanic | 6 (9.0%) |
| Non-Hispanic | 61 (91.0%) |
| Mean Age at Time of Video conference (y) | 10.8 +/- 10.7 |
| Under 18 years of age | 57 (85.1%) |
| Age Range | 3.5 months - 56 years |
| Country of Residence |  |
| USA | 30 (44.8%) |
| United Kingdom | 13 (19.4%) |
| Belgium | 3 (4.5%) |
| Canada | 3 (4.5%) |
| China | 3 (4.5%) |
| Ireland | 3 (4.5%) |
| Brazil | 2 (3.0%) |
| France | 2 (3.0%) |
| Germany | 2 (3.0%) |
| Spain | 2 (3.0%) |
| Australia | 1 (1.5%) |
| Finland | 1 (1.5%) |
| Holland | 1 (1.5%) |
| Poland | 1 (1.5%) |

**Table 2.** *NAA15* demographic information of interviewed patients.

| Characteristics | Study Participants (n=19) n (%) |
| --- | --- |
| Sex |  |
| Male | 14 (73.7%) |
| Female | 5 (26.3%) |
| Race |  |
| Caucasian | 18 (94.7%) |
| Asian | 1 (5.3%) |
| Ethnicity |  |
| Hispanic | 1 (5.3%) |
| Non-Hispanic | 18 (94.7%) |
| Mean Age at Time of Video conference (y) | 8.9 +/- 4.7 |
| Under 18 years of age | 18 (94.7%) |
| Age Range | 2 years - 21 years |
| Country of Residence |  |
| USA | 14 (73.7%) |
| Australia | 3 (15.8%) |
| United Kingdom | 1 (5.3%) |
| Puerto Rico | 1 (5.3%) |

All patients were confirmed to have mutations in the *NAA10* or *NAA15* gene, as shown in **Table 3** and **Table 4**. All individuals were analyzed for pathogenicity using ACMG standards. For probands that possessed novel mutations that have not been characterized in the literature previously, we determined probable pathogenicity based on previous genetic analysis and ACMG standards (see **Supplementary Notes 1-6**) These are denoted with an asterisk (*).

**Table 3.** *NAA10* Variants of Participants. For pathogenicity, P = known pathogenic, LP = likely pathogenic.

| Proband Identifier | Age Range (y) | Sex | Protein Change | cDNA Change | Inheritance | Pathogenicity |
| --- | --- | --- | --- | --- | --- | --- |
| OS_102 | 5-9 | F | Asp10Glu | c.30C>G | De novo | P |
|  |  |  | Pro08Ser | c.22C>T | De novo | P |
| OS_103 | 15-19 | F | His16Pro | c.047A>C | De novo | P |
| OS_104 | 0-4 | F | Tyr31Cys | c.92A>G | De novo | P |
| OS_107 | 55-59 | F | Tyr43Ser | c.128A>C | De novo | P |
| OS_108 | 30-34 | M | Tyr43Ser | c.128A>C | Maternal | P |
| OS_109 | 25-29 | M | Tyr43Ser | c.128A>C | Maternal | P |
| OS_110 | 5-9 | M | Ile072Thr | c.215T>C | Maternal | P |
| OS_111 | 5-9 | M | Ile072Thr | c.215T>C | Maternal | P |
| OS_112 | 5-9 | F | Arg83Cys | c.247C>T | De novo | P |
| OS_113 | 5-9 | F | Arg83Cys | c.247C>T | De novo | P |
| OS_114 | 5-9 | F | Arg83Cys | c.247C>T | De novo | P |
| OS_115 | 10-14 | F | Arg83Cys | c.247C>T | De novo | P |
| OS_116 | 10-14 | F | Arg83Cys | c.247C>T | De novo | P |
| OS_117 | 10-14 | F | Arg83Cys | c.247C>T | De novo | P |
| OS_118 | 35-39 | F | Arg83Cys | c.247C>T | De novo | P |
| OS_119 | 10-14 | F | Arg83Cys | c.247C>T | De novo | P |
| OS_120 | 15-19 | F | Arg83Cys | c.247C>T | De novo | P |
| OS_121 | 10-14 | F | Arg83Cys | c.247C>T | De novo | P |
| OS_122 | 0-4 | F | Arg83Cys | c.247C>T | De novo | P |
| OS_123 | 5-9 | F | Arg83Cys | c.247C>T | De novo | P |
| OS_124 | 15-19 | F | Arg83Cys | c.247C>T | De novo | P |
| OS_125 | 25-29 | F | Arg83Cys | c.247C>T | Maternal | P |
| OS_126 | 0-4 | F | Arg83Cys | c.247C>T | De novo | P |
| OS_127 | 0-4 | F | Arg83Cys | c.247C>T | De novo | P |
| OS_128 | 10-14 | F | Arg83Cys | c.247C>T | De novo | P |
| OS_129 | 5-9 | F | Arg83Cys | c.247C>T | De novo | P |
| OS_130 | 5-9 | F | Arg83Cys | c.247C>T | De novo | P |
| OS_132 | 5-9 | F | Arg83Cys | c.247C>T | De novo | P |
| OS_133 | 0-4 | F | Arg83Cys | c.247C>T | De novo | P |
| OS_135 | 5-9 | F | Arg83Cys | c.247C>T | De novo | P |
| OS_136 | 0-4 | F | Arg83Cys | c.247C>T | De novo | P |
| OS_137 | 0-4 | F | Arg83Cys | c.247C>T | De novo | P |
| OS_138 | 10-14 | M | Arg83Cys | c.247C>T | De novo | P |
| OS_139 | 15-19 | F | Ala087Ser | c.259G>T | Do novo | P |
| OS_140 | 0-4 | F | Ala087Ser | c.259G>T | De novo | P |
| OS_141 | 15-19 | F | Ala87Ser | c.259G>T | De novo | P |
| OS_142 | 10-14 | F | Ala104Asp | c.311C>A | De novo | P |
| OS_143 | 15-19 | F | Arg116Trp | c.346C>T | De novo | P |
| OS_144 | 20-24 | F | His120Pro | c.359A>C | De novo | P |
| OS_146 | 15-19 | F | Leu121Val | c.361C>G | De novo | P |
| OS_147 | 0-4 | F | Ser123Pro | c.367T>C | De novo | P |
| OS_148 | 0-4 | F | Phe128Ser | c.383T>C | De novo | P |
| OS_149 | 0-4 | F | Phe128Leu | c.384T>G | De novo | P |
| OS_150 | 5-9 | F | Phe128Leu | c.384T>G | De novo | P |
| OS_151 | 0-4 | F | Phe128Leu | c.384T>G | De novo | P |
| OS_152 | 5-9 | F | Phe128Leu | c.384T>G | De novo | P |
| OS_153 | 20-24 | F | Phe128Leu | c.384T>G | De novo | P |
| OS_154 | 10-14 | F | Phe128Leu | c.384T>G | De novo | P |
| OS_155 | 0-4 | F | Phe128Leu | c.384T>G | De novo | P |
| OS_156 | 10-14 | F | Met147Thr | c.440T>C | De novo | P |
| OS_157 | 5-9 | F | Met147Thr | c.440T>C | De novo | P |
| OS_159 | 0-4 | M | Thr152Argfs*6 | c.455_458del | Maternal | P |
| OS_160 | 0-4 | M | Glu181Alafs*67 | c.542_551del | De novo | P |
| OS_162 | 10-14 | F | Arg83Cys | c.247C>T | De novo | P |
| OS_163 | 35-39 | F | Met147Thr | c.440T>C | De novo | P |
| OS_164 | 5-9 | M | Arg149Trp | c.445C>T | De novo | P |
| OS_165 | 5-9 | F | Arg116Trp | c.346C>T | De novo | P |
| OS_166 | 0-4 | F | Arg83Cys | c.247C>T | De novo | P |
| OS_167 | 0-4 | F | Arg116Gln | c.347G>A | Unknown | LP* |
| OS_168 | 0-4 | M | Arg116Gln | c347G>A | Maternal | LP* |
| OS_169 | 10-14 | F | Arg116Trp | c.346C>T | Unknown | P |
| OS_170 | 0-4 | M | His34Tyr | c.100C>T | De novo | LP* |
| OS_171 | 20-24 | F | His120Pro | c.359A>C | De novo | P* |
| OS_172 | 5-9 | F | Arg83Cys | c.247C>T | De novo | P |
| OS_181 | 35-39 | F | Met147Leu | c.439A>T | Maternal | LP* |
| OS_187 | 0-4 | F | Met147Leu | c.439A>T | Maternal | LP* |
| OS_188 | 0-4 | M | Met147Leu | c.439A>T | Maternal | LP* |

**Table 4.** *NAA15* Variants of Participants. For pathogenicity, P = known pathogenic, LP = likely pathogenic.

| Proband Identifier | Age Range (y) | Sex | Protein Change | cDNA Change | Inheritance | Pathogenicity |
| --- | --- | --- | --- | --- | --- | --- |
| NAA15_001 | 5-9 | M | p.Arg27Gly (AGA>GGA) | c.79A>G | De novo | P |
| NAA15_002 | 5-9 | M | p.His080Argfs*17 | c.0239_0240del | De novo | P |
| NAA15_003 | 10-14 | M | p.His80Arg fsX17 | c.239_240delAT (frameshift) | De novo | P |
| NAA15_004 | 10-14 | F | p.Trp0083X | c.248G>A | De novo | P |
| NAA15_005 | 0-4 | M | p.Glu173AsnfsX54 | c.517delG | De novo | P |
| NAA15_006 | 5-9 | M | p.Glu173* | c.517G>T | De novo | P |
| NAA15_007 | 5-9 | F | p.L329fs*22 | c.986dupT | De novo | P |
| NAA15_008 | 15-19 | F | p.Thr351Profs*3 | c.1051delA | De novo | P |
| NAA15_009 | 5-9 | M | p.ENST00000296543.5 splice site variant | c.1753+1G>A | Unknown | LP* |
| NAA15_010 | 5-9 | M | p.Asn623Ilefs* | c.1868del | De novo | P |
| NAA15_011 | 20-24 | M | p.Asn0864Ser | c.2591A>G | De novo | P |
| NAA15_012 | 10-14 | M | p.Asp0335Glufs*7 | c.1005-1008del | De novo | P |
| NAA15_013 | 10-14 | F | p.Arg782X | c.2344C>T | De novo | P |
| NAA15_014 | 0-4 | M | p.Ala719Thr (GCA>ACA) | c.2155G>A in exon 17 | De novo | P |
| NAA15_015 | 0-4 | M | p.Ala585Thr | c.1753G>A | De novo | P |
| NAA15_016 | 10-14 | M | p.Ala678Cysfs*3 | c.2031dupT | De novo | P |
| NAA15_017 | 0-4 | M | p.Asn362MetfsX36 | c.1083del | De novo | P |
| NAA15_018 | 5-9 | M | p.Gln0621* | c.1861C>T | De novo | P |
| NAA15_019 | 10-14 | F | p.N/A intronic | c.1087+2T>C | De novo | LP* |

The prevalence of the most common ophthalmic manifestations of OS in our cohort were compared to previously reported cohorts. **Table 5** compares the prevalence of CVI, myopia, astigmatism, strabismus, nystagmus, and hyperopia in our cohort to 106 previously reported cases, for a total of 173 individuals including those reported in this paper. Our cohort had significant overlap with the cohort reported previously^7^. However, the 37 probands from this paper who were not in our cohort were analyzed separately as seen in **Table 5**. Nystagmus and hyperopia were not analyzed in the prior paper, so the prevalence in our cohort was compared to 69 previously reported cases. This data is graphically depicted in **Figure 1**.

**Figure 1.**
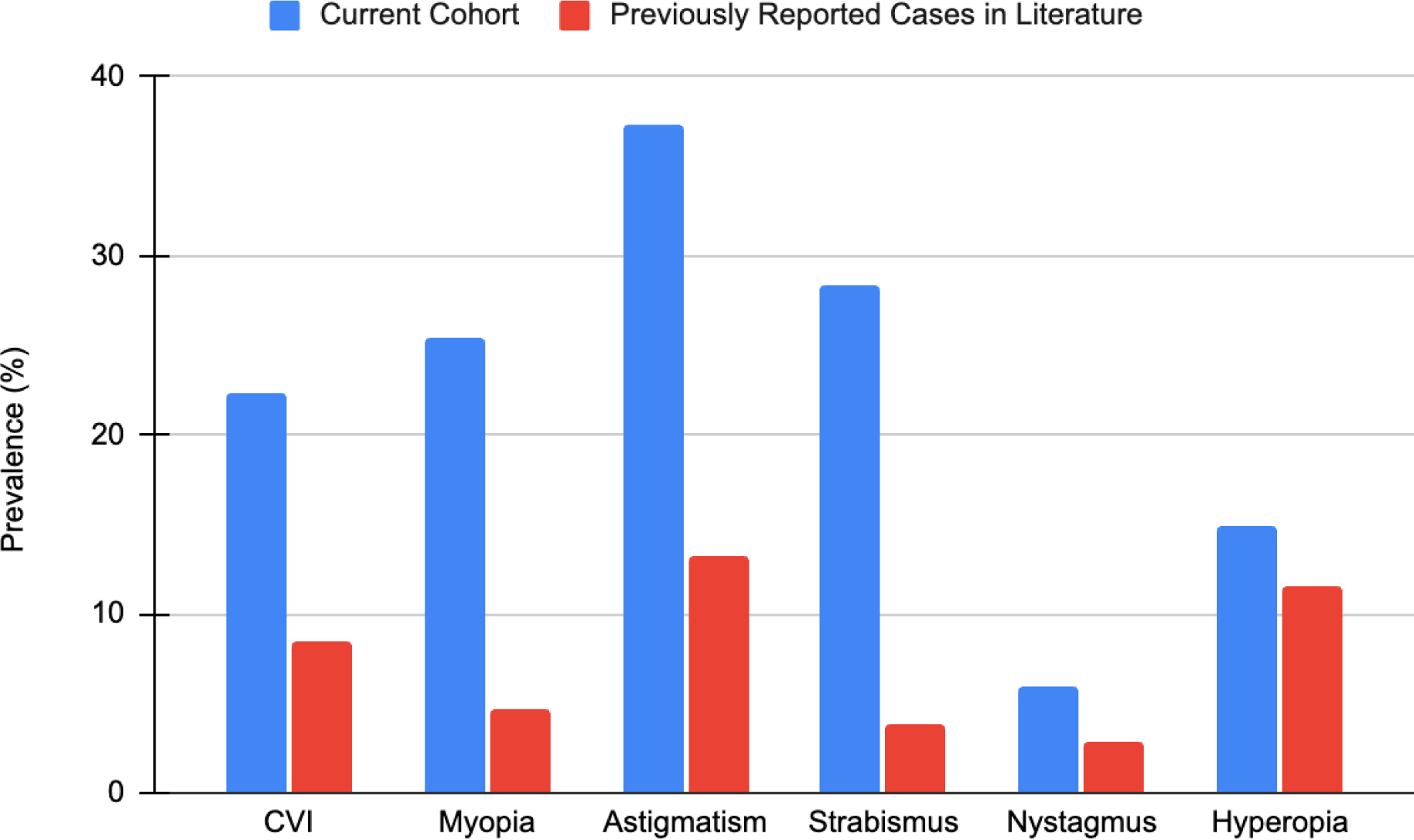
Comparison of Ophthalmic Manifestation Prevalence (%) in Current *NAA10* Cohort versus Previous Literature.

**Table 5.** Comparison of Ophthalmic Manifestations in Current Cohort vs. Previous Literature.

| Condition | Current Cohort (n=67)<br>(%) | Lyon et al., 2023<sup>7</sup>(n=37)<br>(%) | Cheng et al., 2019<sup>15</sup>(n=22)<br>(%) | Gupta et al., 2019<sup>17</sup> (n=39)<br>(%) | McTiernan et al., 2022<sup>6</sup> (n=8)<br>(%) | Weighted total across literature (not including our cohort) | Prevalence including our cohort |
| --- | --- | --- | --- | --- | --- | --- | --- |
| **CVI** | n=15<br>(22.4%) | n=1<br>(2.7%) | n=5<br>(22.7%) | n=3<br>(7.7%) | n=0<br>(0%) | 9/106<br>(8.5%) | 24/173<br>(13.9%) |
| **Myopia** | n=17<br>(25.4%) | n=0<br>(0%) | n=1<br>(4.5%) | n=3<br>(7.7%) | n=1<br>(12.5%) | 5/106<br>(4.7%) | 22/173<br>(12.7%) |
| **Astigmatism** | n=25<br>(37.3%) | n=0<br>(0%) | n=8<br>(36.4%) | n=6<br>(15.4%) | n=0<br>(0%) | 14/106<br>(13.2%) | 39/173<br>(22.5%) |
| **Strabismus** | n=19<br>(28.4%) | n=1<br>(2.7%) | n=2<br>(9.1%) | n=1<br>(2.6%) | n=0<br>(0%) | 4/106<br>(3.8%) | 23/173<br>(13.3%) |
| **Nystagmus** | n=4<br>(6.0%) | Not reported | n=1<br>(4.5%) | n=1<br>(2.6%) | n=0<br>(0%) | 2/69<br>(2.9%) | 6/136<br>(4.4%) |
| **Hyperopia** | n=10<br>(14.9%) | Not reported | n=2<br>(9.1%) | n=4<br>(10.3%) | n=2<br>(25.0%) | 8/69<br>(11.6%) | 18/136<br>(13.2%) |

In comparing the prevalence of CVI, astigmatism, myopia, hyperopia, strabismus, and nystagmus in our *NAA10* versus *NAA15* cohorts, we were unable to ascertain any statistical significance, likely due to small sample size. However, **Figure 2** shows that *NAA10* generally has higher prevalence of ophthalmic manifestations in every formal ophthalmic diagnosis category. Notably, in the absence of a CVI diagnosis, trouble with depth perception was mentioned more frequently in patients with *NAA15* mutations (44.4%, n=8), compared to those with *NAA10* (19.2%, n=1) (**Figure 2**).

**Figure 2.**
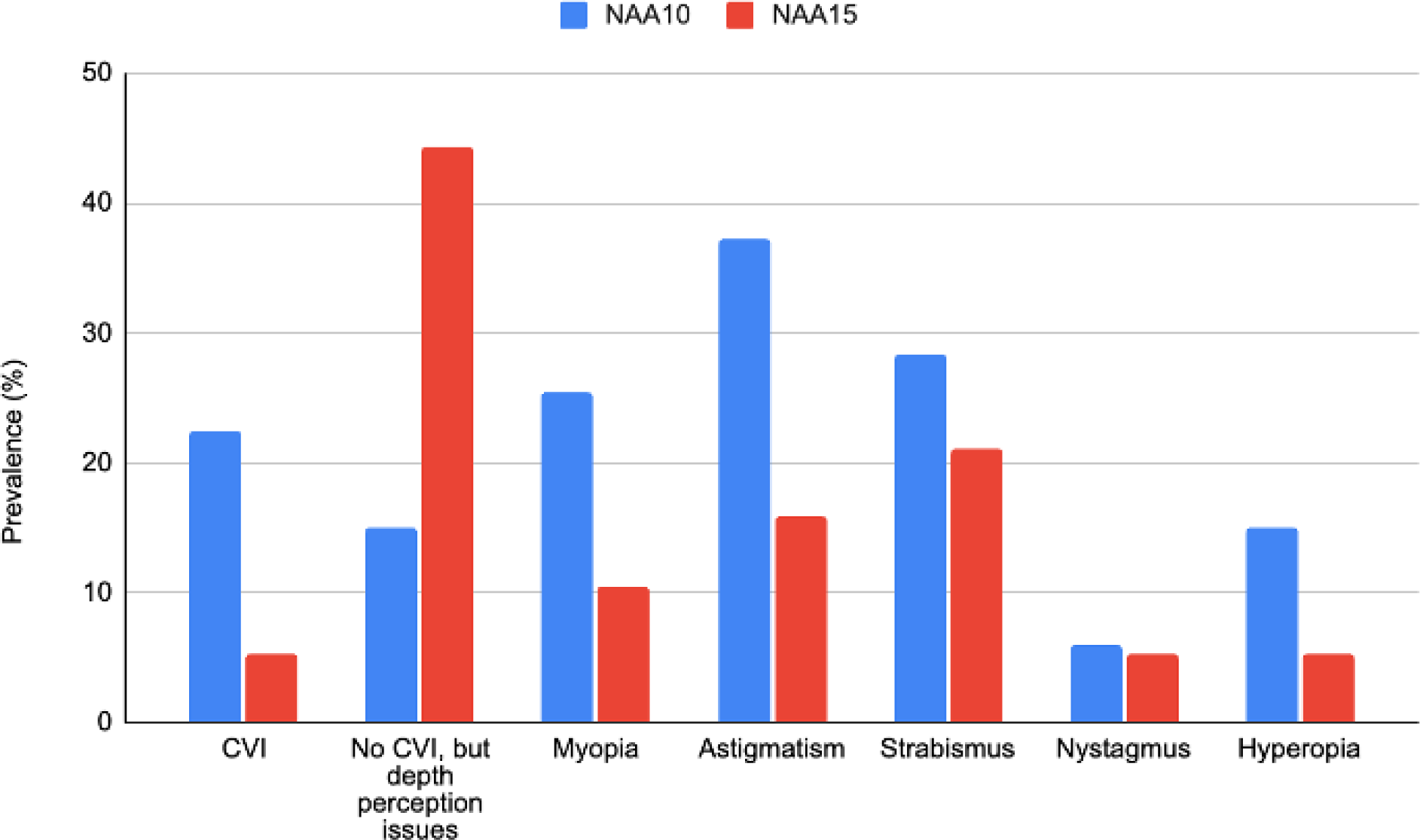
Comparison of ophthalmic manifestation prevalence in *NAA10* versus *NAA15* cohorts

### Neuroimaging

Neuroimaging was analyzed for 13 individuals in the *NAA10* cohort (probands OS_110, OS_111, OS_115, OS_126, OS_132, OS_150, OS_152, OS_153, OS_155, OS_162, OS_163, OS_168, and OS_172) and 5 individuals in the NAA15 cohort (NAA15_007, NAA15_008, NAA15_010, NAA15_013, and NAA15_019) by a neuroradiologist. Probands’ globes were rated 1-4, where 1 is normal, 2 is bilateral dysmorphia, 3 is bilateral dysmorphia and macrophthalmia, and 4 is macrophthalmia, as depicted in **Figure 3**. **Table 6** shows the globe ratings of the 13 assessed *NAA10* probands and **Table 7** shows the globe ratings of the 5 assessed *NAA15* probands, as well as other ophthalmic diagnoses that these patients had at the time of imaging.

**Figure 3.**
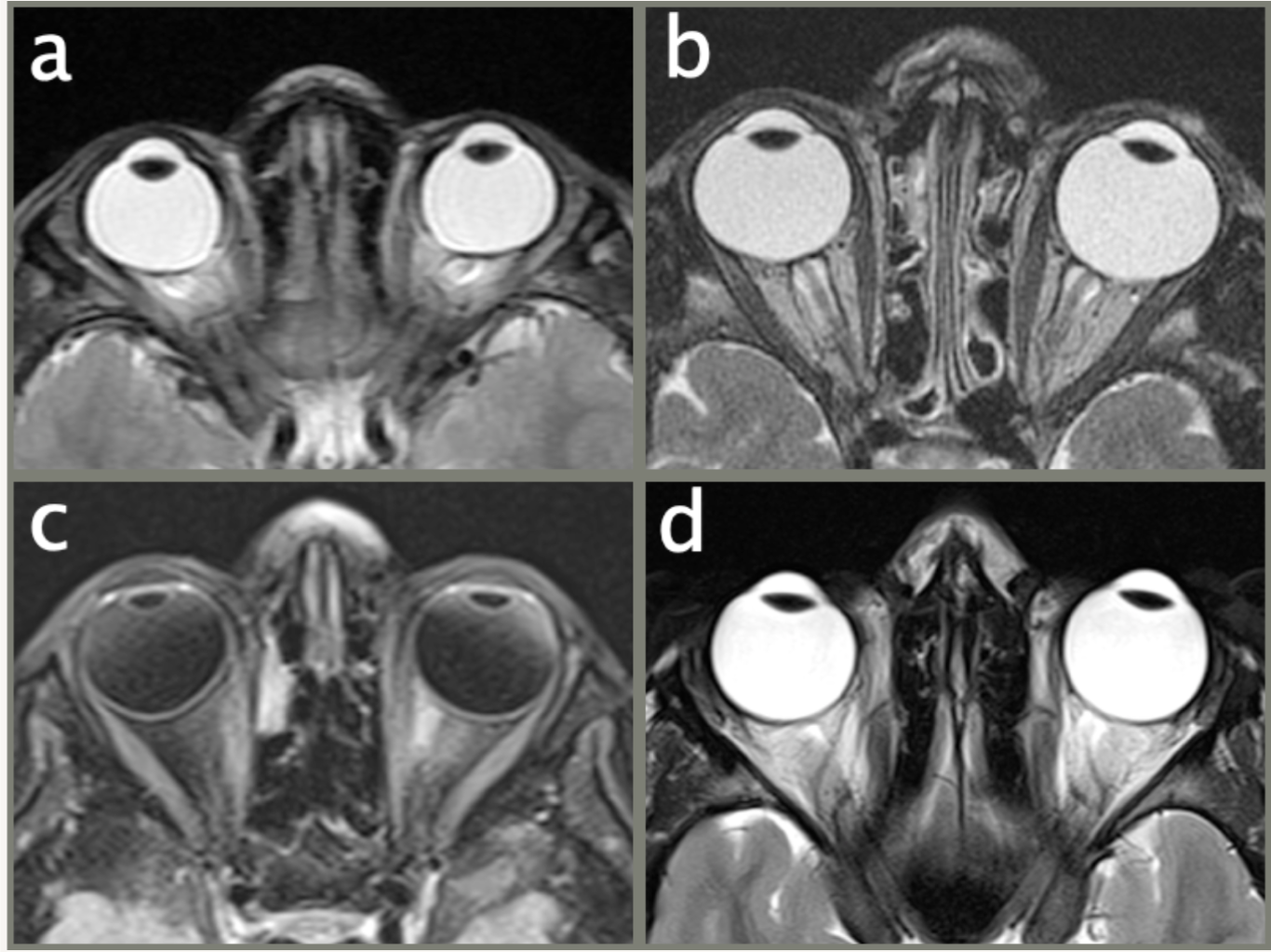
Brain MRI collage depicting ocular globe abnormalities in 4 different patients with Ogden syndrome. (a) axial T2WI (OS_115) shows bilateral globe dysmorphia; (b) axial T2WI (OS_153) shows bilateral macrophthalmia and mild dysmorphia; (c) axial fat-sat T2 FLAIR (OS_150) depicting bilateral macrophthalmia and mild dysmorphia; (d) axial T2WI (patient NAA15_008) shows bilateral macrophthalmia.

**Table 6.** Globe Size and Ophthalmic Diagnoses in *NAA10* Mutation Cohort.

| Proband Identifier | Globe size rating<br>(1-4) | Other Ophthalmology Diagnoses<br>at Time of Imaging |
| --- | --- | --- |
| OS_168 | 1 | None |
| OS_126 | 1 | Astigmatism<br>Strabismus (R Esotropia) |
| OS_172 | 1 | Hyperopia<br>Astigmatism<br>CVI |
| OS_162 | 1 | Myopia<br>CVI<br>Strabismus |
| OS_111 | 1 | None |
| OS_110 | 1 | Strabismus (R esotropia) |
| OS_152 | 1 | Bilateral cortical blindness<br>Astigmatism<br>Myopia<br>Nystagmus<br>Intermittent alternating exotropia |
| OS_155 | 1 | CVI<br>Strabismus<br>Blindness |
| OS_115 (a) | 2 | Myopia<br>CVI |
| OS_132 | 2 | Microphthalmia<br>Optic nerve aplasia<br>Optic nerve hypertrophy<br>CVI<br>Myopia<br>Ptosis |
| OS_163 | 3 | Severe myopia<br>Strabismus (R exotropia) |
| OS_150 (c) | 3 | Nystagmus<br>CVI<br>Oculomotor apraxia<br>Strabismus (intermittent exotropia)<br>Hyperopia |
| OS_153 (b) | 4 | Myopia<br>Strabismus<br>CVI |

**Table 7.** Globe Size and Ophthalmic Diagnoses in *NAA15* Mutation Cohort.

| Proband Identifier | Globe Size Rating (1-4) | Other Ophthalmology Diagnoses at Time of Imaging |
| --- | --- | --- |
| NAA15_007 | 1 | Astigmatism |
| NAA15_010 | 1 | Normal |
| NAA15_013 | 1 | Amblyopia<br>Astigmatism |
| NAA15_019 | 1 | Esotropia |
| NAA15_008 (d) | 4 | Cortical Visual Impairment (CVI)<br>Myopia<br>Pale Optic Nerve<br>Strabismus<br>Nystagmus<br>Progressive Vision Loss<br>Astigmatism |

To better understand the impact of early ophthalmic interventions in patients with Ogden Syndrome, we reached out directly to patient families for quotes (**Table 8**). These underscore the importance of early and regular ophthalmology screening and intervention to optimize mobility, intellectual development, social development, and overall quality of life for patients with *NAA10* and *NAA15* mutations. Full quotes are available in **Supplementary Table S1**.

**Table 8.**
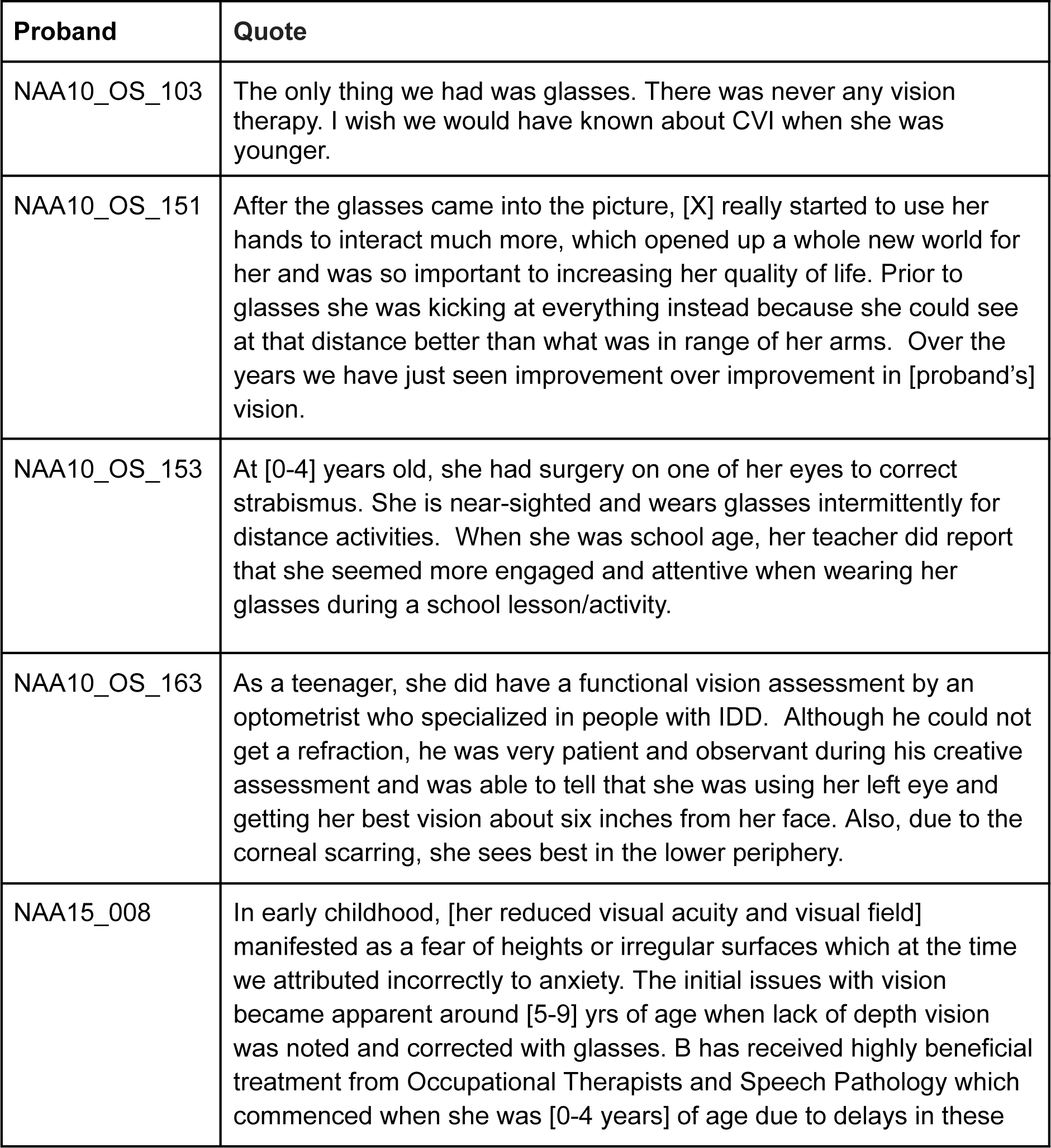
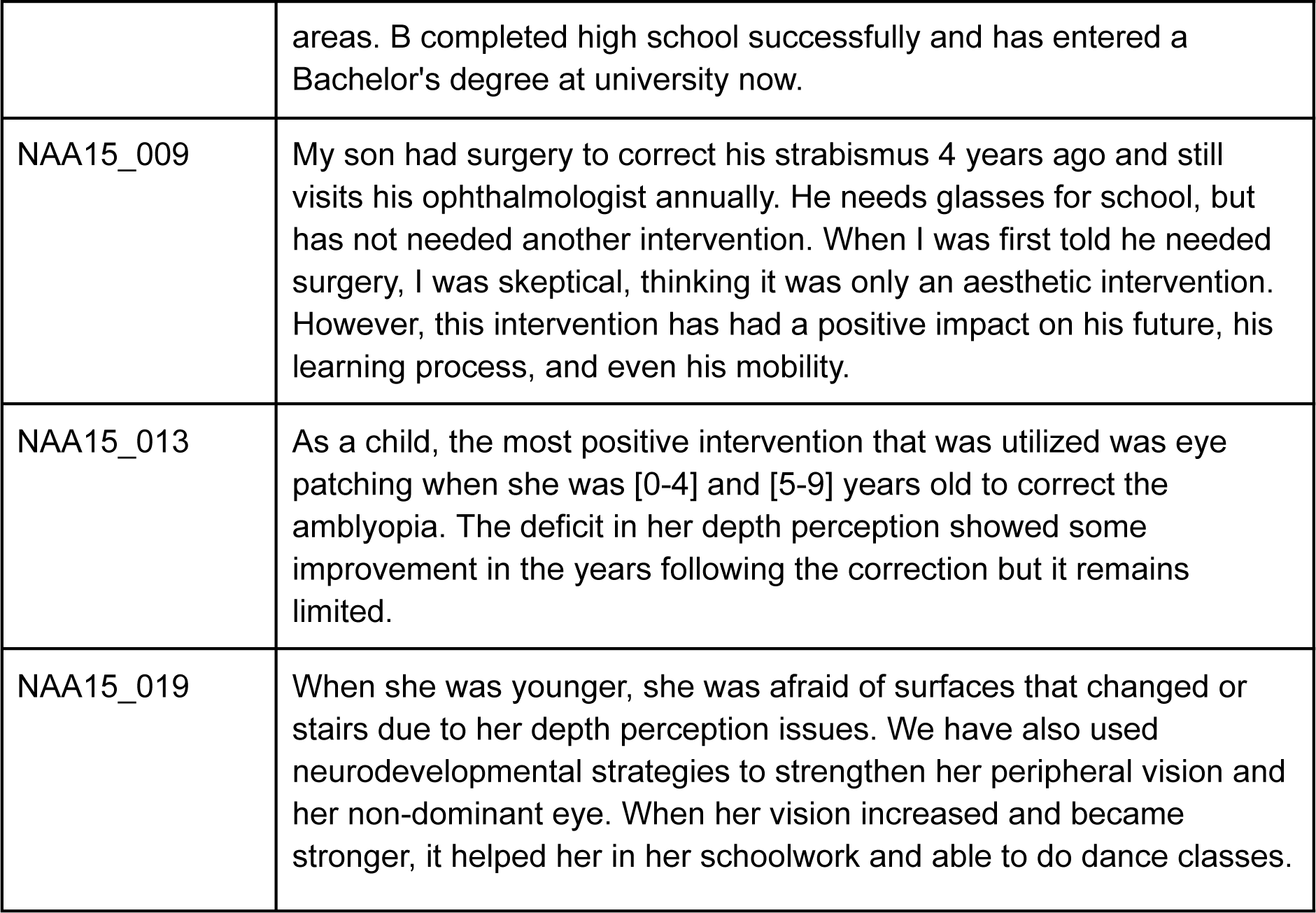
Quotes from *NAA10* and *NAA15* Families.

## Discussion

Mutations in the *NAA10* and *NAA15* genes affect the development of multiple organ systems, including the eyes^11,12,15^. Literature on *NAA10*- and *NAA15*-related neurodevelopmental syndromes to date has not focused on the specifics of ophthalmologic manifestations in a large cohort. We described common ophthalmic conditions in both *NAA10* and *NAA15* individuals including CVI, astigmatism, myopia, hyperopia, strabismus, and nystagmus. The prevalence of all studied conditions was higher in our cohort compared to previous literature (CVI 22.4% vs. 8.5%; myopia 25.4% vs. 4.7%; astigmatism 37.3% vs. 13.2%; strabismus 28.4% vs. 3.8%, respectively). This suggests that many of these conditions may be under-reported and thus underdiagnosed in patients with Ogden Syndrome.

In Western countries, CVI is a leading cause of pediatric bilateral visual impairment^18,19^. It is diagnosed when children exhibit abnormal visual responses that are not caused by the eye itself. As this affects visual processing, it can have far-reaching effects on a patient’s ability to learn and interact with their surroundings. Difficulties with depth perception, face processing, and discernment of contrast are often functional manifestations of CVI^20,21^. It has been suggested that up to 10.5% of children with developmental disabilities have CVI, though many remain undiagnosed^22,23^. Our data support the suggestion that CVI is underdiagnosed, as we found a high prevalence of trouble with depth perception in both *NAA10* and *NAA15* individuals in the absence of a confirmed CVI diagnosis (19.2% (n=1) and 44.4% (n=8), respectively). Further, trouble with depth perception could serve as a metric to better understand visual abnormalities in patients with OS. Examination by an ophthalmologist is recommended upon diagnosis, but further exploration of the characteristics of CVI as it relates to OS could help further guide patient care, ensure early intervention of treatable ocular conditions during development, and provide valuable insights into the underlying mechanisms and development of visual processing.

Previous literature suggests that *NAA10* individuals present with more severe phenotypes than *NAA15* individuals^7^. In terms of the ophthalmic phenotypes, we found no significant differences between *NAA10* and *NAA15* presentations. However, it is important to note the small sample size of the *NAA15* cohort, so this needs to be validated in a larger cohort in the future. Though it was not significant, a higher proportion of *NAA15* patients also had trouble with depth perception with no documented CVI diagnosis. This may be a result of milder phenotypic presentations with *NAA15* mutations, but it may also point to a greater need for screening and assessment for CVI in this patient population.

Interestingly, in comparing presentations across demographics, we found that *NAA10* males were significantly less likely to have astigmatism compared to *NAA10* females (p<0.05) (**Supplementary Figure S1e**). This significance is likely due to a small sample size of males, as mutations in *NAA10* are more lethal in males ^7^. Perhaps *NAA10* males who would have had ophthalmologic manifestations died due to more serious systemic dysfunction. We also analyzed mutation patterns with regards to ophthalmologic manifestations of interest. We found that p.Phe128Leu had a higher prevalence of nystagmus (p<0.05) (**Supplementary Figure S2h**). Again, this may be due to small sample size, but it may also be suggestive of mutation-specific impacts on phenotype. Further research is necessary, as these phenotypic manifestations of specific mutations may aid in guiding screening processes or clinical decision making to improve quality of care for *NAA10* and *NAA15* individuals.

Neuroimaging assessment for structural abnormalites of the ocular globes was performed on a select number of *NAA10* (n=13) and *NAA15* (n=5) individuals. There is no clear correlation between globe size/shape and ocular phenotypes, but it is interesting to note that, overall, the *NAA15* individuals had milder ophthalmic manifestations and fewer neuroimaging abnormalities compared to *NAA10* individuals, with the exception of one *NAA15* individual (p.Thr351Profs*3) who had a more severe ocular presentation than the other *NAA10* individuals. Further neuroimaging and data collection should be done to better understand the functional consequences of ocular globe MRI phenotypes (macrophthalmia and/or dysmorphia) in patients with *NAA10* and *NAA15*-related neurodevelopmental syndromes.

The specific function of *NAA10* and *NAA15* in visual system development are not fully clear. Previous literature has suggested that *NAA10* may be involved in retinoic acid signaling, which is critical for ocular development^12,24^. The critical role of NAA10 in eye development was further supported by a study on zebrafish, wherein NAA10 knockdowns produced small, underdeveloped eyes^25^. *NAA15* has been shown to play a critical role in neuron generation and differentiation^26^, which could also contribute to the visual abnormalities seen in patients with mutations in this protein. Further research is necessary to investigate the connection between *NAA10*/*NAA15* and detrimental alterations in ocular development and function.

## Limitations

A major limitation of this study was that it was conducted through video conferencing and chart review. We were fortunate to have access to thorough medical records for most probands. However, some were not fully comprehensive or did not include records from an ophthalmologist or optometrist. In these instances, we relied on information gathered from interviews with the patients’ parents, who may not always recall details of medical information. As a result of this, we are likely under-reporting the prevalence of the ophthalmic manifestations studied.

Additionally, our cohorts contained some novel mutations, which made it difficult to classify some proband mutations as pathogenic versus likely pathogenic versus VUS. However, this was greatly aided by recently published material on *NAA10* and *NAA15* mutation pathogenicity^7^. Further, there are mutations, such as p.Ser123Pro, that have been previously published as “likely pathogenic,” but biochemical analysis has not been performed. We used ACMG criteria by Richards et al. to classify mutations that have not been reported or classified previously^27^. These classification justifications are available in **Supplementary Notes S(1-6).**

## Conclusions

*NAA10*-related and *NAA15*-related neurodevelopmental syndrome is classically characterized by intellectual disability, congenital cardiac anomalies, and delayed development. However, our analysis underscores the importance of considering the ophthalmic phenotypes that are common in this condition. The prevalence of CVI, myopia, astigmatism, strabismus, nystagmus, and hyperopia were higher in our cohort of 67 patients with *NAA10* mutations than previously reported in literature. To our knowledge, this is the first cohort analysis of the ocular manifestations of *NAA15* mutations. Our analysis suggests that many of these conditions are likely being underdiagnosed in this patient population, specifically CVI. More research with larger cohorts and functional assays is certainly needed to better understand the role that these mutations play in altering visual development. This would help promote prompt screening, assessment, and intervention by an optometrist or ophthalmologist upon diagnosis of Ogden syndrome and improve the quality of life for these patients.

## Data Availability

All data produced in the present study are available upon reasonable request to the authors, except for private identifiable patient information.

## Author Contributions

GJL conducted all virtual interviews with participants and was responsible for primary data collection, with assistance by EM. RP was responsible for data analysis and project conception, along with GJL. MW was responsible for neuroimaging analysis. The first draft of the manuscript was written by RP and AP, with critical revision performed by GJL, AG, and MW at several points.

## Acknowledgments

We thank the families for their participation and support.

## Ethical Approval

Both oral and written patient consent were obtained for research and publication, with approval of protocol #7659 for the Jervis Clinic by the New York State Psychiatric Institute - Columbia University Department of Psychiatry Institutional Review Board.

## Supplementary Information

**Figures S1(a-h).** Prevalence of the Ophthalmic Manifestations of NAA10 Mutations by Demographic (n=67).

**Figures S2(a-h).** Prevalence of the Ophthalmic Manifestations of NAA10 Mutations by Mutation (n=50). Note that only mutations with ≥ 3 probands were included in the analysis.

**Table S1.** Full quotes from select *NAA10* and *NAA15* families.

**Notes S(1–6).** NAA10 Mutations - Arg116Gln, His120Pro, His034Tyr, Met147Leu. NAA15 Mutation - p.ENST00000296543.5 splice site variant c.1753+1G>A

**Supplementary Dataset S1.** Excel file: NAA10 Summary

**Supplementary Dataset S2.** Excel file: NAA15 Summary

