## Supplementary Information for "Ophthalmic Manifestations of NAA10-Related and NAA15-Related Neurodevelopmental Syndrome: Analysis of Cortical Visual Impairment and Refractive Errors"

### Supplementary Data

**Figures S1(a-h).** Prevalence of the Ophthalmic Manifestations of NAA10 Mutations by Demographic (n=67).

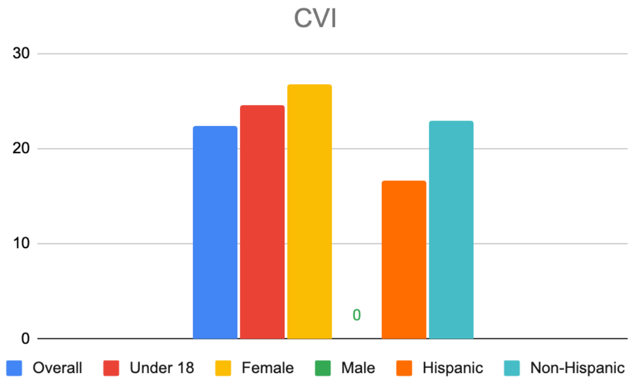


**Figure S1a.** Prevalence (%) of CVI by Demographic in Patients with NAA10 Mutations (n=67).

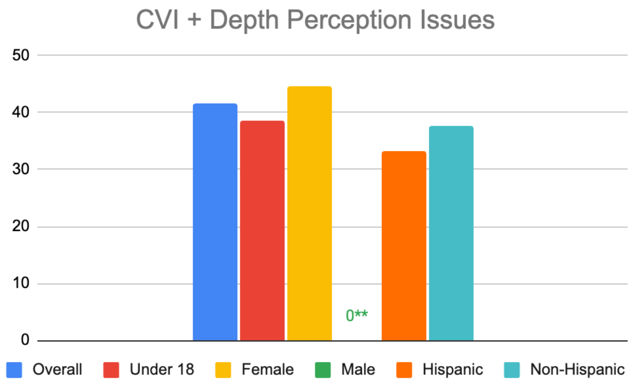


**Figure S1b.** Combined Prevalence (%) of CVI and Depth Perception Issues by Demographic in Patients with NAA10 Mutations (n=67). **p<0.01

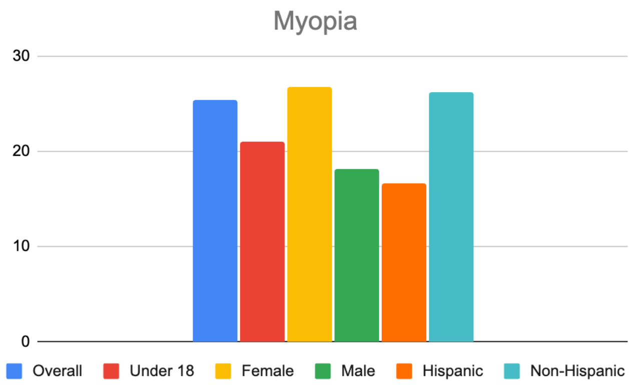


**Figure S1c.** Prevalence (%) of Myopia by Demographic in Patients with NAA10 Mutations (n=67).

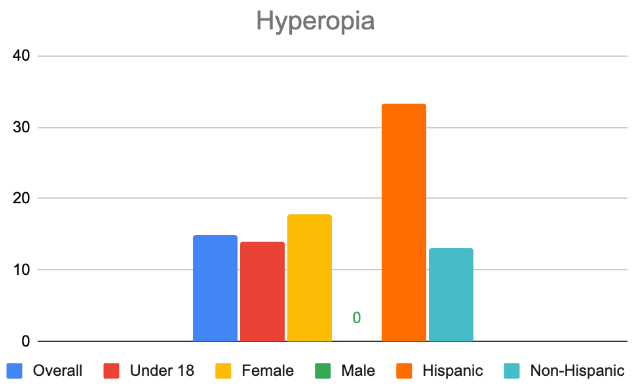


**Figure S1d.** Prevalence (%) of Hyperopia by Demographic in Patients with NAA10 Mutations (n=67).

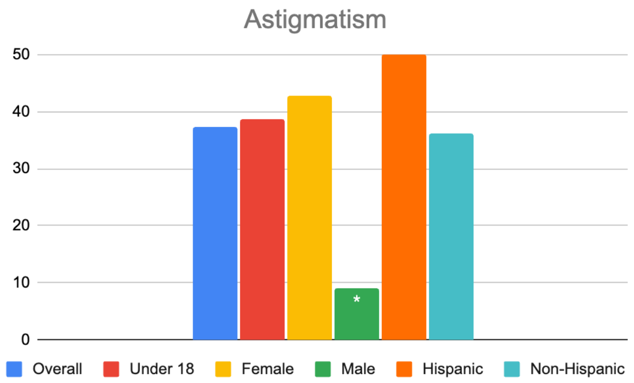


**Figure S1e.** Prevalence (%) of Astigmatism by Demographic in Patients with NAA10 Mutations (n=67). *p<0.05

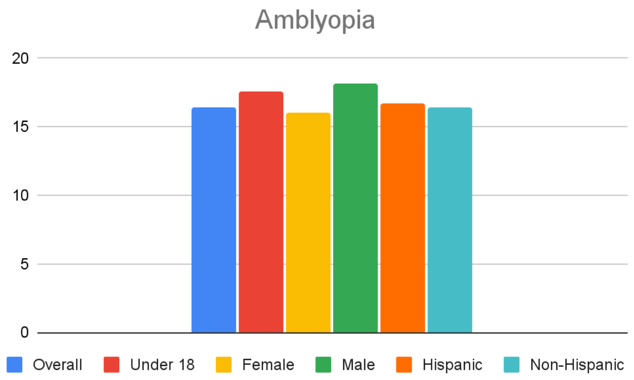


**Figure S1f.** Prevalence (%) of Amblyopia by Demographic in Patients with NAA10 Mutations (n=67).

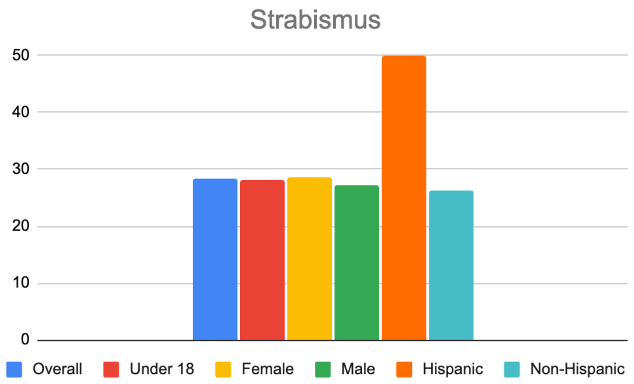


**Figure S1g.** Prevalence (%) of Strabismus by Demographic in Patients with NAA10 Mutations (n=67).

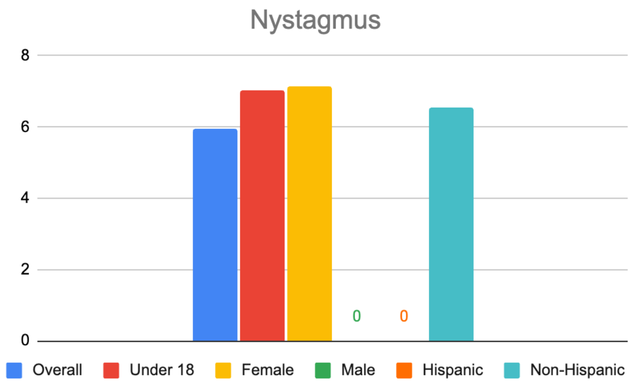


**Figure S1h.** Prevalence (%) of Nystagmus by Demographic in Patients with NAA10 Mutations (n=67).

**Figures S2(a-h).** Prevalence of the Ophthalmic Manifestations of NAA10 Mutations by Mutation (n=50). Note that only mutations with ≥ 3 probands were included in the analysis.

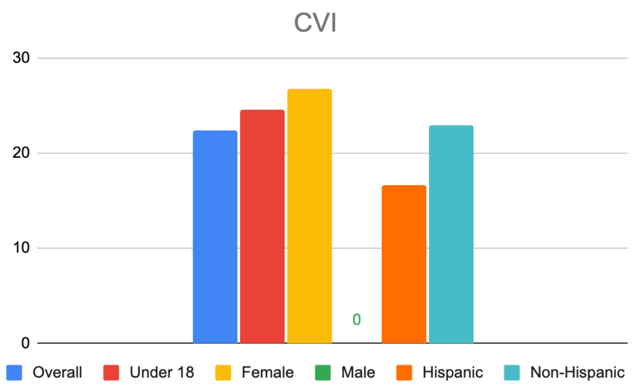


**Figure S2a.** Prevalence (%) of CVI by Mutation in Patients with NAA10 Mutations (n=50).

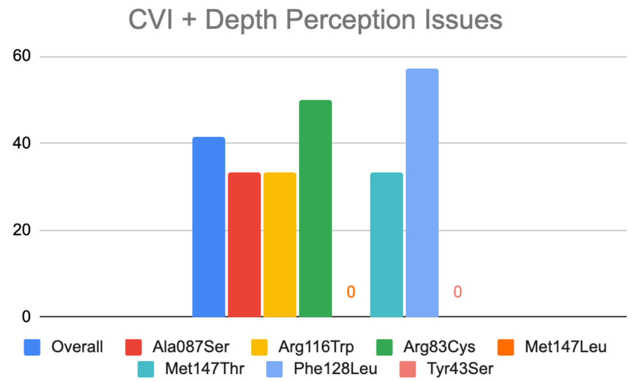


**Figure S2b.** Combined Prevalence (%) of CVI and Depth Perception Issues by Mutation in Patients with NAA10 Mutations (n=50).

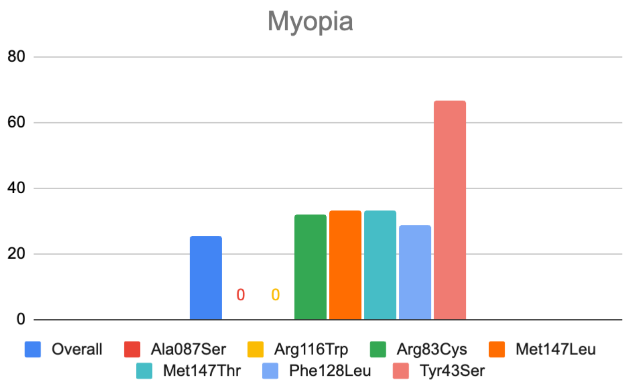


**Figure S2c.** Prevalence (%) of Myopia by Mutation in Patients with NAA10 Mutations (n=50).

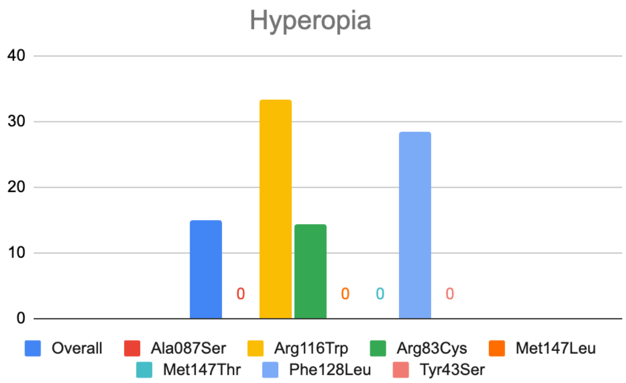


**Figure S2d.** Prevalence (%) of Hyperopia by Mutation in Patients with NAA10 Mutations (n=50).

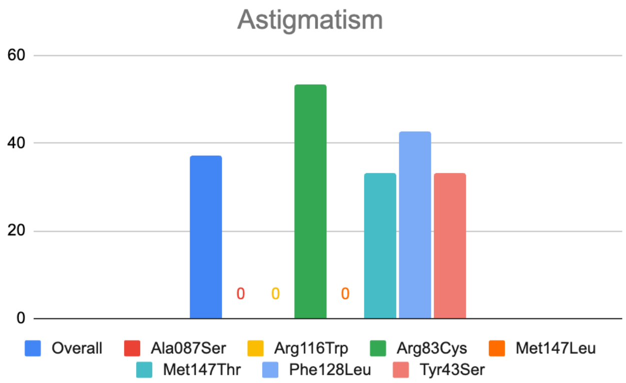


**Figure S2e.** Prevalence (%) of Astigmatism by Mutation in Patients with NAA10 Mutations (n=50).

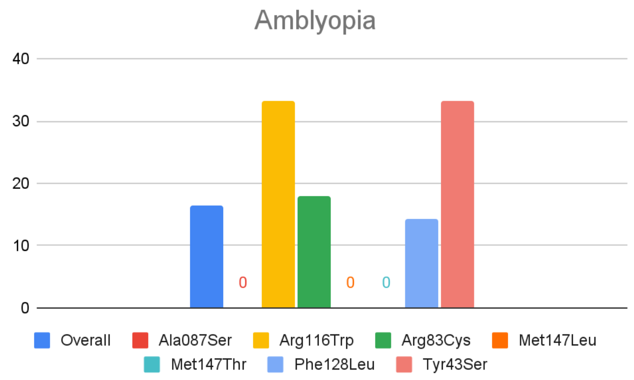


**Figure S2f.** Prevalence (%) of Amblyopia by Mutation in Patients with NAA10 Mutations (n=50).

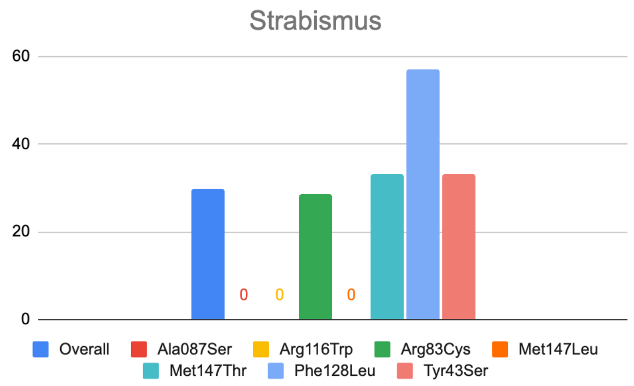


**Figure S2g.** Prevalence (%) of Strabismus by Mutation in Patients with NAA10 Mutations (n=50).

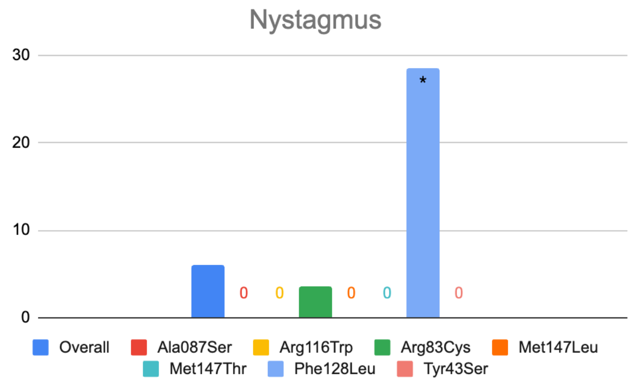


**Figure S2h.** Prevalence (%) of Nystagmus by Mutation in Patients with NAA10 Mutations (n=50). *p<0.05

**Table S1.** Full quotes from select *NAA10* and *NAA15* families.

| Proband | Full Quote |
|---|---|
| NAA10_OS_103 | The only thing we had was glasses. There was never any vision therapy. I wish we would have known about cvi when she was younger. I believe she has it but if I hunted down the diagnosis there really isn't anything that would help her being she is [20-24 years] old.. |
| NAA10_OS_151 | Soon after [X] was born and we began noticing she wasn't visually attending as she should, we took her to her first Ophthalmology appointment and were very abruptly told [X] was blind and dismissed out of the room. There was no further guidance from this particular Doctor and we left that appointment feeling just absolutely devastated and confused. A couple months passed and we noticed [X] swatting at brightly colored and backlit games on our phone and realized she wasn't blind at all. [X] entered early intervention programs at [0-4 years] old where we were introduced to a vision therapist who was able to introduce the idea of CVI/neurological blindness. Later with a diagnosis at [0-4 years], we booked an intensive therapy appointment in Arizona for [X] with a woman named Michelle Turner who runs a program called "Movement Lesson" to help children with mobility issues. Michelle emphasized the importance of a visual diagnosis to maximize the effectiveness of her program so she recommended a clinic in Phoenix that specialized in CVI. It was there that the Ophthalmologist told us [X] was extremely farsighted and would benefit from glasses. After the glasses came into the picture [X] really started to use her hands to interact much more which opened up a whole new world for her and was so important to increasing her quality of life. Prior to glasses she was kicking at everything instead because she could see at that distance better than what was in range of her arms.<br>Over the years we have just seen improvement over improvement in [X]'s vision. We can not express enough the significance of finding the right Ophthalmologist for children with Ogden Syndrome or getting an early diagnosis so families of these children can be advised on this topic by the other families that have already been through it. We can't imagine where we would have been if we would have just went with the clinical picture that was painted for us by the first vision specialist [X] was seen by. To go from being told your child is blind at [0-4 years] old to now understand at [0-4] years old [X] is visually impaired but her brain is learning how to see is such a monumental shift and we can't imagine the position of families left without proper guidance indefinitely.<br>We also highly recommend families add a vision therapist to their therapy rotation. Thankfully [X] was put in contact with one early on through the Early Intervention Program and continues to be followed by one at her Developmental Preschool but if families have not yet been in contact with one they should heavily consider seeking one out. They can help with specific recommendations to help someone struggling with CVI learn to see better such as using high contrast or lights to the persons advantage. |

| | |
|---|---|
| NAA10_OS_153 | Our daughter, [X], is [20-24] years old and has Ogden Syndrome. She has been seen by Ophthalmologists annually from infancy to the present. The main observation has always been that she has a "pale optic nerve" and consequently Cortical Vision Impairment (CVI). Also, at [0-4] years old, she had surgery on one of her eyes to correct strabismus.<br>[X] is severely cognitively and physically disabled. She is non-verbal, non-ambulatory and requires maximum assistance with all activities of daily living. Assessing her vision is difficult and cannot be done by all Ophthalmologists/Optometrists, in our experience. Finding the right practitioner who understands the challenges of assessing vision of a severely disabled person is key. She has worn glasses over the years (from approximately age [10-14] until present). She is near-sighted and wears glasses intermittently for distance activities. When she was school age, her teacher did report that she seemed more engaged and attentive when wearing her glasses during a school lesson/activity.<br>Working with a Vision Therapist has been very helpful over the years. They have ensured that she was presented things on a black or white background to provide contrast for her to see things better. |
| NAA10_OS_163 | When she was [0-4 years] old she saw an ophthalmologist who noted intermittent right eye exotropia (strabismus). He said it was so intermittent that we should not worry. The next time she saw an ophthalmologist she was six or seven years old. By that time, she had lost vision in her right eye having developed amblyopia. He said that we should have had the strabismus corrected when she was young. Of course, in hindsight, we wish we had had her followed up sooner. At the same time (age [5-9]) she had pretty severe myopia in both eyes and depth perception issues from the amblyopia in the right eye. However, getting an accurate refraction was difficult due to her severe photophobia and sensory defensiveness around her face. She would not tolerate glasses (except for a brief period of about six months at age [5-9], in spite of repeated trials with new refractions. By age [5-9] or so, her myopia was so severe that she was assessed as legally blind and received vision specialist services through school. At about age [15-19] she had an episode of corneal hydrops in her right eye and was diagnosed with keratoconus, likely explaining why eyeglass corrections were never helpful as the shape of her cornea was likely changing frequently. Later, around age [20-24] she had another hydrops episode in her left eye. While she was recovering from that, she was essentially completely blind -- proving that she had little to no vision in the right eye. She did have an ophthalmologic exam under anesthesia (the only way to adequately assess the health of her eyes). But for her developmental issues (severe IDD, photophobia and constant eye rubbing) she would be a candidate for corneal transplant, but she would not be able to tolerate the aftercare. We had this opinion from three different ophthalmologists. She also had developed a cataract in the right eye, likely due to trauma of constant rubbing. Sometime in the last few years (age 35-39) her lens in that eye has become completely detached, falling into the vitreous. As a teenager, she did have a functional vision assessment by an optometrist who specialized in people with IDD particularly difficult to assess. Although he could not get a refraction, he was very patient and |

| | |
|---|---|
| | observant during his creative assessment and was able to tell that she was using her left eye and getting her best vision about six inches from her face. Also, due to the corneal scarring, she sees best in the lower periphery. |
| NAA10_OS_170 | Fortunately [X] has no ophthalmology issues to date. We were seeing ophthalmology 2x per year, but now cut back to 1x per year. |
| NAA15_008 | [X] has nystagmus and reduced visual acuity and visual field which became apparent prior to her diagnosis with NAA15 related disorder in her mid-teens. In early childhood this manifested as a fear of heights or irregular surfaces which at the time we attributed incorrectly to anxiety. The initial issues with vision became apparent around [5-9] yrs of age when lack of depth vision was noted and corrected with glasses. [X] has received highly beneficial treatment from Occupational Therapists and Speech Pathology which commenced when she was [0-4 years] of age due to delays in these areas. Seeking appropriate supports at school and accommodations has been a continual challenge and ultimately required changes of school in both primary and secondary school. [X] completed high school successfully and has entered a Bachelor's degree at university now. |
| NAA15_009 | We have a child with NAA15 syndrome. His name is [X] and we live in Puerto Rico. He was part of Dr. Lyon's research. He was intervened for strabismus on October, 2019. He is now [10-14] years old and still visits his ophthalmologist annually. He needs glasses for school, but has not needed another intervention.<br>My interest in sharing this is in part because I think it's important for families to be aware of the risks these kids face when going to surgery. Fortunately, we had a geneticist who was available to help with the sedation process and was consulted on the safest options for him. At the time, he was also taking a carnitine supplement that he no longer uses.<br>When I was first told he needed the surgery, I was skeptical, thinking it was only an esthetic intervention. It was vital for me to have a doctor who could explain the importance of this intervention for his future, his learning process and even his mobility. |
| NAA15_013 | As a child, the most positive intervention that was utilized was eye patching when she was [0-4] and [5-9] years old. It helped to correct the amblyopia. The deficit in her depth perception showed some improvement in the years following the correction but it remains limited.<br>I wish that I could provide some positive feedback regarding her visual development. Unfortunately, [X]'s vision has been a significant issue in recent years. She has recently been diagnosed with low vision and she is struggling with adapting to the changes. We have seen a decrease in her vision over the past 5 years. It is complicated as she has difficulty describing her difficulties and exams have been challenging. She seems to struggle more with reading text. At this point, we are working on the adaptive changes that are now needed for her diagnosis but, obviously, we wish for clearer answers as to how and why this has happened. |

| | |
|---|---|
| NAA15_019 | Good afternoon, [X] is c.1087+2T>C in the article. [X] is extremely far sighted with exotropia. She also has an astigmatism and diminished depth perception. Becca received therapies since age [0-4 years] and received her glasses at age [0-4]. I believe the early intervention of glasses and therapies including PT and OT that did address her depth perception and visual impairments has helped her as she is now age [10-14]. She can track words and read. When she was younger she was afraid of surfaces that changed or stairs due to her depth perception issues. We have also used neuro developmental strategies to strengthen her peripheral vision and her non dominant eye. When her vision increased and became stronger it helped her in her schoolwork and able to do dance classes. She continues to receive eye exams by a pediatric ophthalmologist twice a year and retinal eye mapping once a year. |

**Note S(1-6).** Mutation classification justifications for mutations that were not previously classified or reported.

**Note S1.** *NAA10* Mutation: Arg116Gln

(S1a) Criteria 1: PS3 (Well established in vitro or in vivo functional studies supportive of a damaging effect on the gene or gene product)

Justification: "The two identified de novo missense variants [**Arg116Trp** and Val107Phe] in the catalytic N-acetyltransferase domain of *Naa10* **affect highly conserved amino-acid residues both in orthologous and paralogous genes**. As in silico predictions using different algorithms classified both variants between benign and deleterious, we further studied them using 3D homology modeling. These **protein structure-based predictions revealed that the Trp116 mutation most probably hampers CoA binding and reduces the enzymatic activity of Naa10**" (Popp et al., 2014)

(S1b) Criteria 2: PM2 (Absent from controls (or at extremely low frequency if recessive) in Exome Sequencing Project, 1000 Genomes Project, or Exome Aggregation Consortium)

Justification: Absent from gnomAD (Exome Aggregation Consortium)

(S1c) Criteria 3: PM5 (Novel missense change at an amino acid residue where a different missense change determined to be pathogenic has been seen before. Example: Arg156His is pathogenic; now you observe Arg156Cys)

Justification: Arg116Trp shown to be pathogenic by Lyon et al., 2023 and Popp et al., 2014

(S1d) Criteria 4: PP3 (Multiple lines of computational evidence support a deleterious effect on the gene or gene product (conservation, evolutionary, splicing impact, etc.))

Justification: Adapted from Popp et al., 2014 supplementary table 1

| AS Change | SIFT score | Prediction | PolyPhen Score | Prediction | PANTHER score | $P_{deleterious}$ | SNAP Accuracy | Prediction |
|---|---|---|---|---|---|---|---|---|
| p.Arg116Trp | 0,00 | Affects protein function | 0,187 | Benign | -4.95 | 0.875 | 60% | neutral |

(S1e) Classification: **LIKELY PATHOGENIC** (iii)

1 Strong (PS3) AND 1-2 Moderate (PM2, PM5)

**Note S2.** *NAA10* Mutation: His120Pro

(S2a) Criteria 1: PM2 (Absent from controls (or at extremely low frequency if recessive) in Exome Sequencing Project, 1000 Genomes Project, or Exome Aggregation Consortium)

Justification: Absent from gnomAD

(S2b) Criteria 2: PS3 (Well established in vitro or in vivo functional studies supportive of a damaging effect on the gene or gene product)

Justification: As reported in the paper by Lyon et al., 2023: "Based on these structures, the *NAA10* variants p.Pro8Ser/Asp10Glu, p.Tyr31Cys, **p.His120Pro**, p.Ser123Pro, p.Phe128Ser, and p.Arg149Trp **would likely compromise the folding and/or thermal**

**stability of NAA10.** Specifically, the mutations to proline (p.His120Pro, p.Ser123Pro) occur in helices, likely destabilizing the helical structure."

(S2c) Criteria 3: PS2 (De novo (both maternity and paternity confirmed) in a patient with the disease and no family history)

Justification: Confirmed de novo mutation via genomic sequencing of patient and both parents.

(S2d) Classification: **PATHOGENIC (ii)**

Greater than or equal to 2 Strong (PS2, PS3)

**Note S3.** *NAA10* Mutation: His034Tyr

(S3a) Criteria 1: PM2 (Absent from controls (or at extremely low frequency if recessive) in Exome Sequencing Project, 1000 Genomes Project, or Exome Aggregation Consortium)

Justification: Absent from gnomAD

(S3b) Criteria 2: PS2 (De novo (both maternity and paternity confirmed) in a patient with the disease and no family history

Justification: Confirmed de novo mutation via exome sequencing study. Patient has no family history of this condition.

(S3c) Classification: **LIKELY PATHOGENIC (ii)**

1 strong (PS2) AND 1-2 Moderate (PM2)

*note: exome sequencing study in this patient confirmed "likely pathogenic" classification

**Note S4.** *NAA10* Mutation: Met147Leu

(S4a) Criteria 1: PM2 (Absent from controls (or at extremely low frequency if recessive) in Exome Sequencing Project, 1000 Genomes Project, or Exome Aggregation Consortium)

Justification: Absent from gnomAD

(S4b) Criteria 2: PP1 (Cosegregation with disease in multiple affected family members in a gene definitively known to cause the disease)

Justification: This mutation has not been previously published, but appeared in a mother and her 2 children

(S4c) Criteria 3: PP4 (Patient's phenotype or family history is highly specific for a disease with a single genetic etiology)

Justification: The affected mother and her 2 children have the same genetic mutation with similar phenotypes. Mutations in NAA10 are specific for Ogden's Syndrome.

(S4d) Criteria 4: PM5 (Novel missense change at an amino acid residue where a different missense change determined to be pathogenic has been seen before. Example: Arg156His is pathogenic; now you observe Arg156Cys)

Justification: Met147Thr was shown to be pathogenic previously by Lyon et al., 2023

(S4e) Criteria 5: PS3 (Well established in vitro or in vivo functional studies supportive of a damaging effect on the gene or gene product)

Justification: As shown in the Cheng et al. (2019) paper, there is a well established functional assay demonstrating that Met147Thr had an effect on enzymatic activity and thermal stability of the NatA complex. This particular allele (Met147Leu) has not been directly tested in this assay, so it is debatable whether PS3 applies or not in this instance.

(S4f) Classification: **Likely pathogenic (v), not including PS3 criteria**

2 Moderate (PM2, PM5) AND greater than or equal to 2 supporting (PP1, PP4)

Note: this is not including PS3 criteria because this is debatable. If PS3 is included, this would constitute a pathogenic (ii) mutation with 2 strong criteria met.

**Note S5**. *NAA15* Mutation: p.ENST00000296543.5 splice site variant c.1753+1G>A

(S5a) Criteria 1: PVS1 (null variant (nonsense, frameshift, canonical +/- 1 or 2 splice sites, initiation codon, single or multiexon deletion) in a gene where LOF is a known mechanism of disease

Justification: This variant is a change of guanine in the invariant GT donor splice site for exon 14 to an adenine, which is predicted to result in aberrant exon splicing. Cheng et al. (2018) reported that loss of function in *NAA15* are associated with varying levels of intellectual disability, autism spectrum disorder, and congenital anomalies.

(S5b) Criteria 2: PM6 (Assumed de novo, but without confirmation of paternity and maternity)

Justification: Likely de novo, parents not tested. Genetic testing summary (summary said inheritance “unknown”)

(S5c) Classification: **LIKELY PATHOGENIC (i)**

1 very strong (PVS1) AND 1 moderate (PM6)

*note: likely pathogenic mutation confirmed by genetic sequencing results

**Note S6.** NAA15 Mutation: p.N/A intronic, c1087+2T>C

(S6a) Criteria 1: PS2 (De novo (both maternity and paternity confirmed) in a patient with the disease and no family history)

Justification: Confirmed de novo. Mother and father do not have the mutation. No family history.

(S6b) Criteria 2: PM2 (Absent from controls (or at extremely low frequency if recessive) in Exome Sequencing Project, 1000 Genomes Project, or Exome Aggregation Consortium)

Justification: absent from gnomAD

(S6c) Classification: **Likely pathogenic (ii)**

1 strong (PS2) AND 1 moderate (PM2)
